# Changes in Health Behaviors and Correlational Patterns among High School Students Across Two Longitudinal Studies

**DOI:** 10.64898/2026.11.17.26368320

**Authors:** Shan Li, Elyse Ganss, Cristian Gomez, Adam Lazarchak, Connor M. Moriarty, Russell K. McIntire

## Abstract

**Purpose:** This study examines whether six health behaviors remain stable among high school students across the school year and whether associations between these behaviors recur across assessments.

**Design/methodology/approach:** As part of a school health program evaluation, a pre-post longitudinal health survey was conducted among two samples of high school students in a U.S. vocational-technical high school. Study 1 surveyed 114 students from late winter to late spring in 2024. Study 2 surveyed 177 students from autumn 2024 to late spring 2025. Wilcoxon signed-rank tests examined group-level behavioral change, and Pearson correlation networks described associations among six health behaviors: sleep, breakfast consumption, physical activity, nature exposure, social media use, and mindfulness practices.

**Findings:** The primary analyses detected no significant group-level changes in any of the six behaviors across either study interval, suggesting relative stability within these samples. A one-time survey may provide useful information for broad school health planning, whereas repeated assessment remains necessary for evaluating longer-term patterns or program effects. Sleep and breakfast showed the most consistent positive association. Physical activity and nature exposure were positively associated at three of the four assessments. Social media was isolated from the other behaviors at the first three assessments but was negatively associated with sleep and physical activity at the final assessment.

**Originality:** The study distinguishes stability in group-level behavior from changes in the relationships among behaviors. The findings support efficient assessment for broad educational planning. They also identify behavior pairs that can be addressed together in school health education, including sleep and breakfast, physical activity and nature exposure, and social media, sleep, and physical activity.

**Research type:** Research paper

## 1. Introduction

Health behaviors established during adolescence often persist into adulthood and significantly influence long-term health trajectories (Harris, 2010; Tamanal and Kim, 2020). Although extensive research has examined adolescents’ sleep, diet, physical activity, social media use, and stress management habits, the potential influence of seasonal variation remains a methodological factor that is often overlooked or underexplored in behavioral studies. For instance, prior evidence on whether seasonal shifts meaningfully influence children’s physical activity remains mixed. Rich et al. (2012) found that UK-based studies consistently reported higher activity in summer and lower in winter, whereas several non-UK studies detected no seasonal differences. Among Danish children, objectively measured moderate-to-vigorous physical activity was 23% lower in winter than in spring (Hjorth *et al*., 2013). The same study also found that children slept approximately 2% longer during winter compared to spring, indicating modest seasonal variation in sleep duration. Health educators designing school-based interventions therefore need solid evidence about which behaviors reflect stable patterns and which fluctuate naturally over time.

Research examining multiple health behaviors simultaneously has increasingly adopted network analysis approaches to understand how behaviors cluster and influence one another. Rather than treating behaviors as independent targets, network perspectives recognize that changes in one behavior may ripple through connected behaviors (Hevey, 2018). Physical activity and time spent in nature, for instance, frequently co-occur; outdoor time is strongly associated with increased physical activity levels among youth, and this relationship may mediate positive mental health outcomes (Bélanger *et al*., 2019). Similarly, sleep duration predicts breakfast consumption among adolescents (Lebacq *et al*., 2022). A micro-longitudinal study found that poorer sleep on a given night reduced the likelihood of eating breakfast the following morning (Mathew *et al*., 2022). Identifying which behavioral connections remain stable provides insight into behavioral clusters that interventions might leverage.

This study addresses two research questions among vocational-technical high school students. First, do reported health behaviors show significant change across assessment intervals spanning different portions of the school year? Second, do the correlational patterns characterizing relationships between these behaviors recur across assessments and studies? To address these questions, the present study focuses on six core health behaviors commonly examined in adolescent health research: sleep duration, breakfast consumption, physical activity, nature exposure, social media use, and mindfulness practice. We conducted two longitudinal studies at the same vocational-technical high school. Study 1 examined a winter to spring interval within one semester. Study 2 extended the investigation to a subsequent school year collection spanning autumn to spring. We applied the same analytical approaches and decisions to both studies. The studies are reported separately, with an integrated interpretation. If health behaviors and their interrelationships vary across these intervals, single-timepoint assessments may capture only transient patterns. Conversely, recurring findings across assessments may identify patterns that warrant further investigation as potential intervention targets.

## 2. Measures and Analytical Approach

### 2.1 Research context and ethical procedures

During February–May 2024, a vocational-technical high school in the Northeastern United States hosted an Intro to Well-Being program administered by a behavioral health services organization. The program consisted of between one and four in-class sessions designed to develop students’ self-awareness, sense of belonging and connection, and reduce stress. To evaluate the impact of this program on the health of students, researchers conducted pre-and-post surveys among intervention and comparison classrooms, both of which were selected based on teachers’ voluntary interest in the program and/or data collection. A more detailed description of the program is found in a previous study (Bucher et al., 2025). Subsequently, the program and evaluation, with similar data collection instruments and methods, were again conducted during the following school-year yet expanded to begin September 2024 and conclude in May 2025. Data were collected through structured questionnaires administered in-class during the school day. The current analysis focuses on health behavior variables that were included in the original program evaluation studies as covariates, not as the primary outcomes. Additionally, as a precursor to the current analysis, we checked whether exposure to the Intro to Well-Being program influenced the strength of correlation between these health behaviors. We computed partial correlations for each key edge on the pooled sample, controlling for group (1 = comparison, 2 = intervention). We did not find any significant differences by intervention and comparison groups, so we feel confident pooling these groups within each survey administration, respectively. These findings provide reassurance that the descriptive association patterns were not substantially altered by adjustment for group assignment. Institutional ethical approval was obtained before data collection, and both studies followed the same consent procedures.

### 2.2 Health Behaviors

Participants self-reported six health behaviors using a structured questionnaire. Sleep duration was assessed by asking, “On an average school night, how many hours of sleep do you get?”. The response options range from *4 or fewer hours* to *10 or more hours*, coded on a 7-point scale (1 = 4 or fewer hours; 7 = 10 or more hours). Breakfast consumption was measured by the number of days participants ate breakfast during the past week. The options ranged from *0 day* to *7 days* and were coded on an 8-point scale (1 = 0 days; 8 = 7 days). Physical activity was assessed by asking how many days in the past week participants engaged in at least 60 minutes of activity that increased their heart rate and breathing. Responses were coded on an 8-point scale, ranging from *0* to *7 days*.

Nature exposure was measured by asking participants to estimate weekly hours spent in outdoor activities (e.g., walking, gardening, camping). Responses ranged from *1*–*3 hours* to *15 or more hours* and were coded on a 5-point scale (1 = 1–3 hours; 5 = 15 or more hours). Social media use was measured by frequency of use, ranging from *I do not use social media* to *More than once an hour*, coded on an 8-point scale (1 = none; 8 = more than once an hour). Mindfulness practice was assessed by asking how often participants engaged in techniques such as meditation, yoga, or breathing exercises. Responses were recorded on a 7-point scale, where 1 indicated ‘*never’* and 7 indicated ‘*more than once a day*.’

### 2.3 Data Analyses

For both studies, Time 1 denotes the initial assessment and Time 2 denotes the follow-up assessment. The elapsed interval differed by study: approximately three months in Study 1 and eight months in Study 2. All within-study analyses compared Time 1 with Time 2. To examine whether individual health behaviors demonstrate stability in their central tendency across time, normality assumptions for paired-samples t-tests were first evaluated for each of the six health variables. Assessment of difference scores (Time 2 - Time 1) using Shapiro-Wilk tests revealed violations of normality assumptions for all six behaviors in both studies. Therefore, Wilcoxon signed-rank tests were conducted instead. For each comparison, we also calculated a rank-biserial correlation as an effect-size measure. The p-value indicates the strength of evidence against the null hypothesis of no significant change, whereas the rank-biserial correlation describes the direction and magnitude of the paired difference. Positive rank-biserial values indicate generally higher responses at Time 2, and negative values indicate generally lower responses at Time 2.

Moreover, contemporaneous networks were constructed separately for each measurement time point to examine the cross-sectional relationships among health behavior variables at each wave. The networks represent the correlational structure of studying variables, where nodes represent variables and edges represent correlations. The networks provide insight into how different health behaviors cluster together at each time point. To ensure meaningful interpretation and avoid including trivial relationships, a significance threshold was applied to determine which edges to include in the networks. Correlations with absolute values greater than 0.2 were considered significant and included as edges in the network visualization. This threshold was selected based on conventional guidelines for meaningful effect sizes in psychological research, which consider correlations of 0.2 to be small to moderate (Gignac and Szodorai, 2016). Edges meeting the significance criteria were represented as solid lines with thickness proportional to the absolute correlation strength. For comparative purposes in the Time 2 network, relationships that were significant at Time 1 but fell below the threshold at Time 2 were included as dashed lines to facilitate direct comparison of network changes.

## 3 Study 1

### 3.1 Participants and procedure

For study 1, we recruited 114 participants from the vocational-technical high school. Of the participants, 39 were 15 years old, 48 were 16 years old, and 27 were 17 years old or older. There were 74 male students, 37 female students, 1 student who identified as non-binary/third gender, and 2 students who did not disclose their gender. Data collection occurred at two timepoints: February 26–28, 2024 (late winter) and May 23–29, 2024 (late spring). This three-month interval captured the seasonal transition while maintaining the same academic semester context. After removing cases lacking complete data, analyses included 113 participants at the first timepoint and 108 at the second. Paired samples for individual behaviors ranged from 111 to 113.

### 3.2 Results

The primary paired analyses did not reveal significant changes in any behavior (Table 1). Unadjusted *p*-values were .673 for sleep, .829 for breakfast, .865 for physical activity, .621 for social media, .563 for mindfulness practices, and .053 for nature exposure. Nature exposure showed the largest increase (rank-biserial *r* = .297; Table 2). However, after the Holm correction accounted for testing all six behaviors, the adjusted *p*-value increased to .319 from .053. The remaining Holm-adjusted *p*-values were 1.000.

**Table 1.**
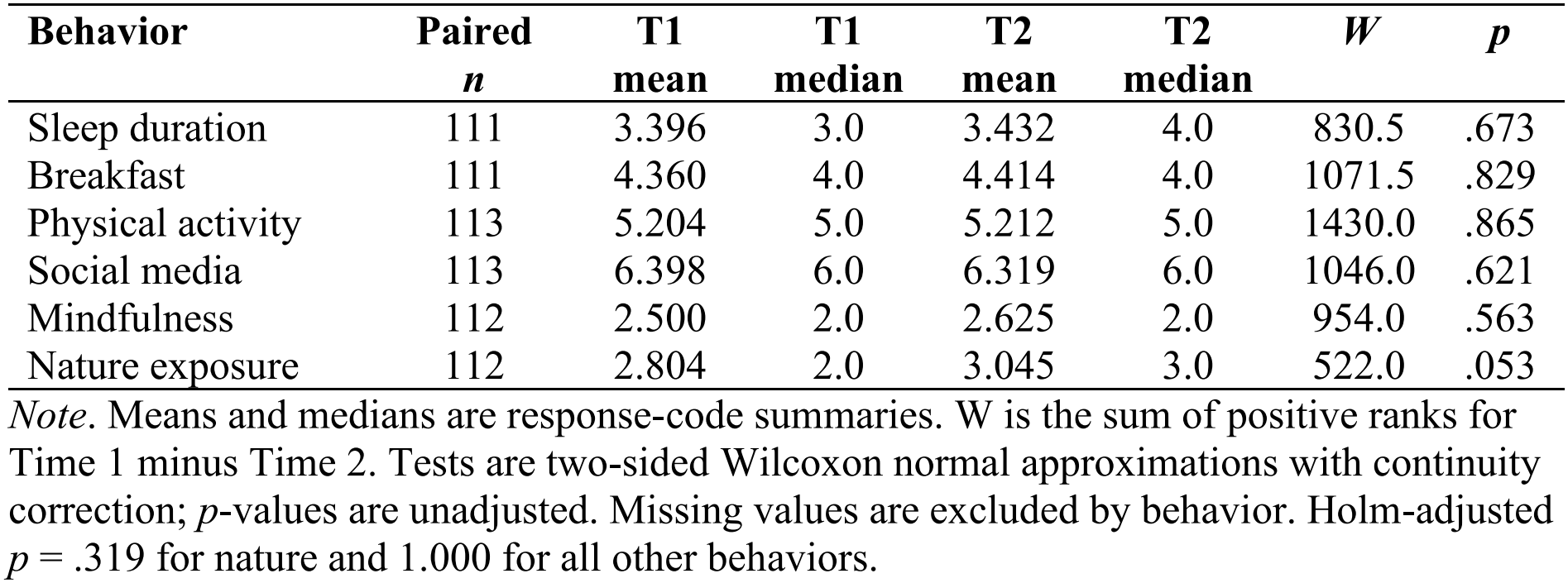
Paired health behavior comparisons in Study 1.

| **Behavior** | **Paired *n*** | **T1 mean** | **T1 median** | **T2 mean** | **T2 median** | ***W*** | ***p*** |
|---|---|---|---|---|---|---|---|
| Sleep duration | 111 | 3.396 | 3.0 | 3.432 | 4.0 | 830.5 | .673 |
| Breakfast | 111 | 4.360 | 4.0 | 4.414 | 4.0 | 1071.5 | .829 |
| Physical activity | 113 | 5.204 | 5.0 | 5.212 | 5.0 | 1430.0 | .865 |
| Social media | 113 | 6.398 | 6.0 | 6.319 | 6.0 | 1046.0 | .621 |
| Mindfulness | 112 | 2.500 | 2.0 | 2.625 | 2.0 | 954.0 | .563 |
| Nature exposure | 112 | 2.804 | 2.0 | 3.045 | 3.0 | 522.0 | .053 |

*Note*. Means and medians are response-code summaries. W is the sum of positive ranks for Time 1 minus Time 2. Tests are two-sided Wilcoxon normal approximations with continuity correction; *p*-values are unadjusted. Missing values are excluded by behavior. Holm-adjusted *p* = .319 for nature and 1.000 for all other behaviors.

**Table 2.**
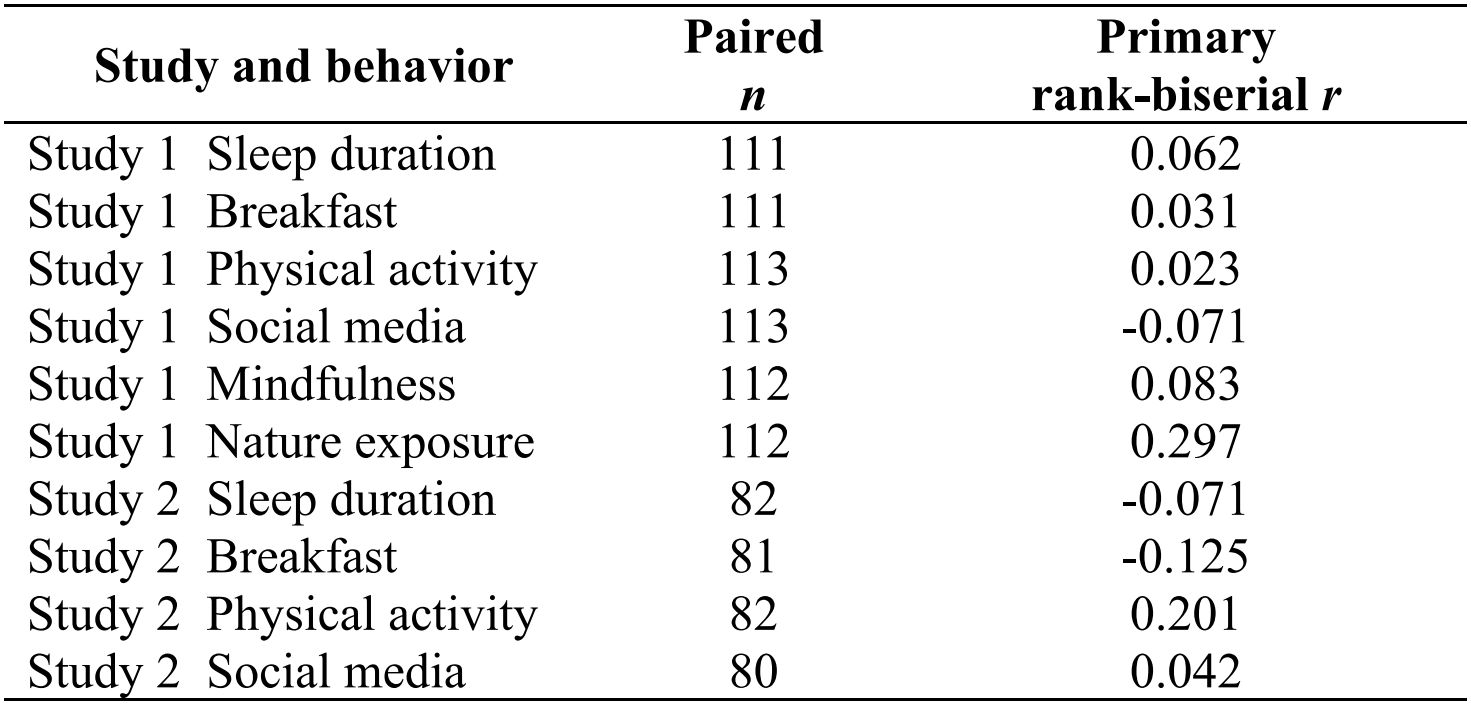
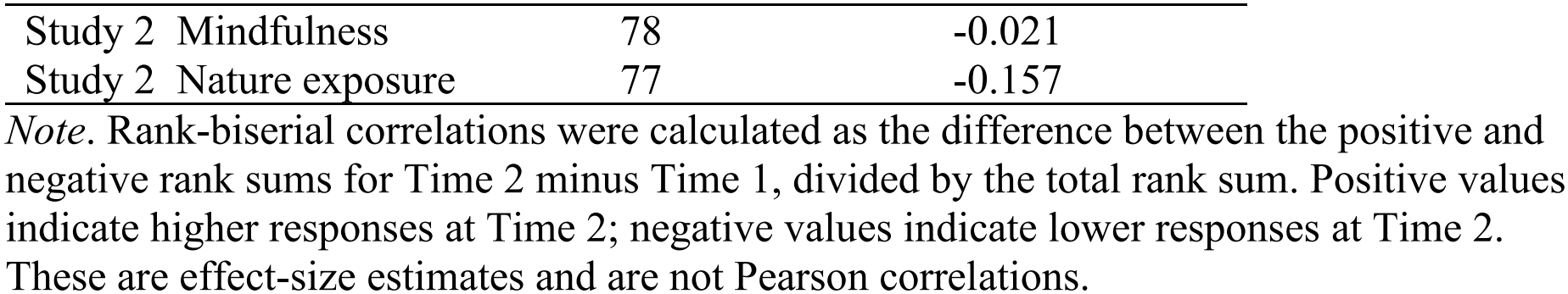
Primary rank-biserial effect sizes across both studies.

| **Study and behavior** |  | **Paired**<br>***n*** | **Primary**<br>**rank-biserial *r*** |
|---|---|---|---|
| Study 1 | Sleep duration | 111 | 0.062 |
| Study 1 | Breakfast | 111 | 0.031 |
| Study 1 | Physical activity | 113 | 0.023 |
| Study 1 | Social media | 113 | -0.071 |
| Study 1 | Mindfulness | 112 | 0.083 |
| Study 1 | Nature exposure | 112 | 0.297 |
| Study 2 | Sleep duration | 82 | -0.071 |
| Study 2 | Breakfast | 81 | -0.125 |
| Study 2 | Physical activity | 82 | 0.201 |
| Study 2 | Social media | 80 | 0.042 |
| Study 2 | Mindfulness | 78 | -0.021 |
| Study 2 | Nature exposure | 77 | -0.157 |

*Note*. Rank-biserial correlations were calculated as the difference between the positive and negative rank sums for Time 2 minus Time 1, divided by the total rank sum. Positive values indicate higher responses at Time 2; negative values indicate lower responses at Time 2. These are effect-size estimates and are not Pearson correlations.

Table 3 reports all 15 pairwise Pearson correlations at each assessment. Table 4 summarizes three prominent pairs in Study 1: physical activity-nature, sleep-breakfast, and breakfast-mindfulness. Physical activity-nature correlations were .383 and .393, and sleep-breakfast correlations were .379 and .323, at the two assessments (Figure 1; Table 4). Breakfast-mindfulness declined from .238 to .103 and appeared as a dashed edge at Time 2. Social media had no connection above the .20 display threshold at either wave.

**Table 3.** Full Pearson correlations.

| **Pair** | **Study 1 T1** | **Study 1 T2** | **Study 2 T1** | **Study 2 T2** |
|---|---|---|---|---|
| Sleep duration-Breakfast | 0.379 | 0.323 | 0.193 | 0.324 |
| Sleep duration-Physical activity | -0.046 | -0.016 | 0.063 | 0.072 |
| Sleep duration-Social media | 0.051 | 0.035 | -0.022 | -0.209 |
| Sleep duration-Mindfulness | 0.075 | 0.121 | -0.015 | 0.069 |
| Sleep duration-Nature exposure | 0.020 | 0.054 | -0.065 | 0.117 |
| Breakfast-Physical activity | -0.076 | 0.107 | -0.022 | 0.038 |
| Breakfast-Social media | -0.080 | -0.079 | -0.162 | 0.014 |
| Breakfast-Mindfulness | 0.238 | 0.103 | 0.104 | 0.024 |
| Breakfast-Nature exposure | 0.020 | -0.043 | 0.000 | 0.096 |
| Physical activity-Social media | -0.038 | -0.035 | 0.006 | -0.238 |
| Physical activity-Mindfulness | 0.081 | 0.078 | 0.003 | -0.051 |
| Physical activity-Nature exposure | 0.383 | 0.393 | 0.038 | 0.268 |
| Social media-Mindfulness | 0.083 | 0.037 | 0.023 | 0.032 |
| Social media-Nature exposure | -0.009 | -0.043 | 0.124 | -0.187 |
| Mindfulness-Nature exposure | 0.096 | 0.102 | -0.019 | -0.276 |

*Note*. Complete-case samples: Study 1, n = 113/108; Study 2, n = 170/77.

**Table 4.**
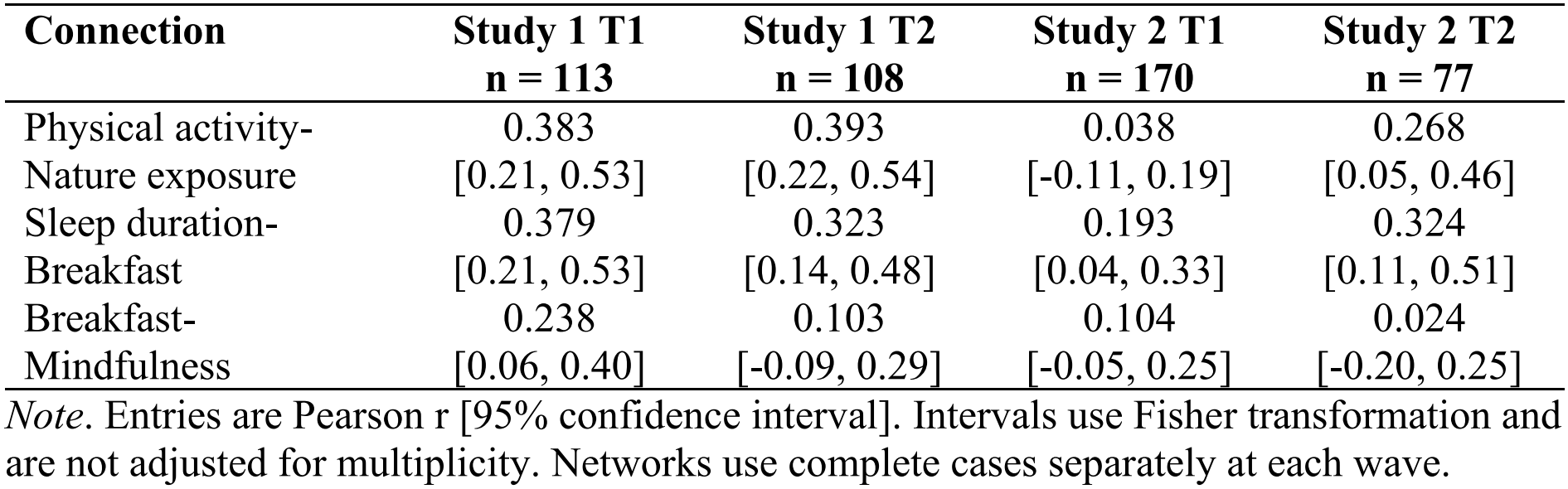
Focal Pearson correlations across the two studies.

| **Connection** | **Study 1 T1**<br>**n = 113** | **Study 1 T2**<br>**n = 108** | **Study 2 T1**<br>**n = 170** | **Study 2 T2**<br>**n = 77** |
|---|---|---|---|---|
| Physical activity-Nature exposure | 0.383<br>[0.21, 0.53] | 0.393<br>[0.22, 0.54] | 0.038<br>[-0.11, 0.19] | 0.268<br>[0.05, 0.46] |
| Sleep duration-Breakfast | 0.379<br>[0.21, 0.53] | 0.323<br>[0.14, 0.48] | 0.193<br>[0.04, 0.33] | 0.324<br>[0.11, 0.51] |
| Breakfast-Mindfulness | 0.238<br>[0.06, 0.40] | 0.103<br>[-0.09, 0.29] | 0.104<br>[-0.05, 0.25] | 0.024<br>[-0.20, 0.25] |

*Note*. Entries are Pearson r [95% confidence interval]. Intervals use Fisher transformation and are not adjusted for multiplicity. Networks use complete cases separately at each wave.

**Figure 1.**
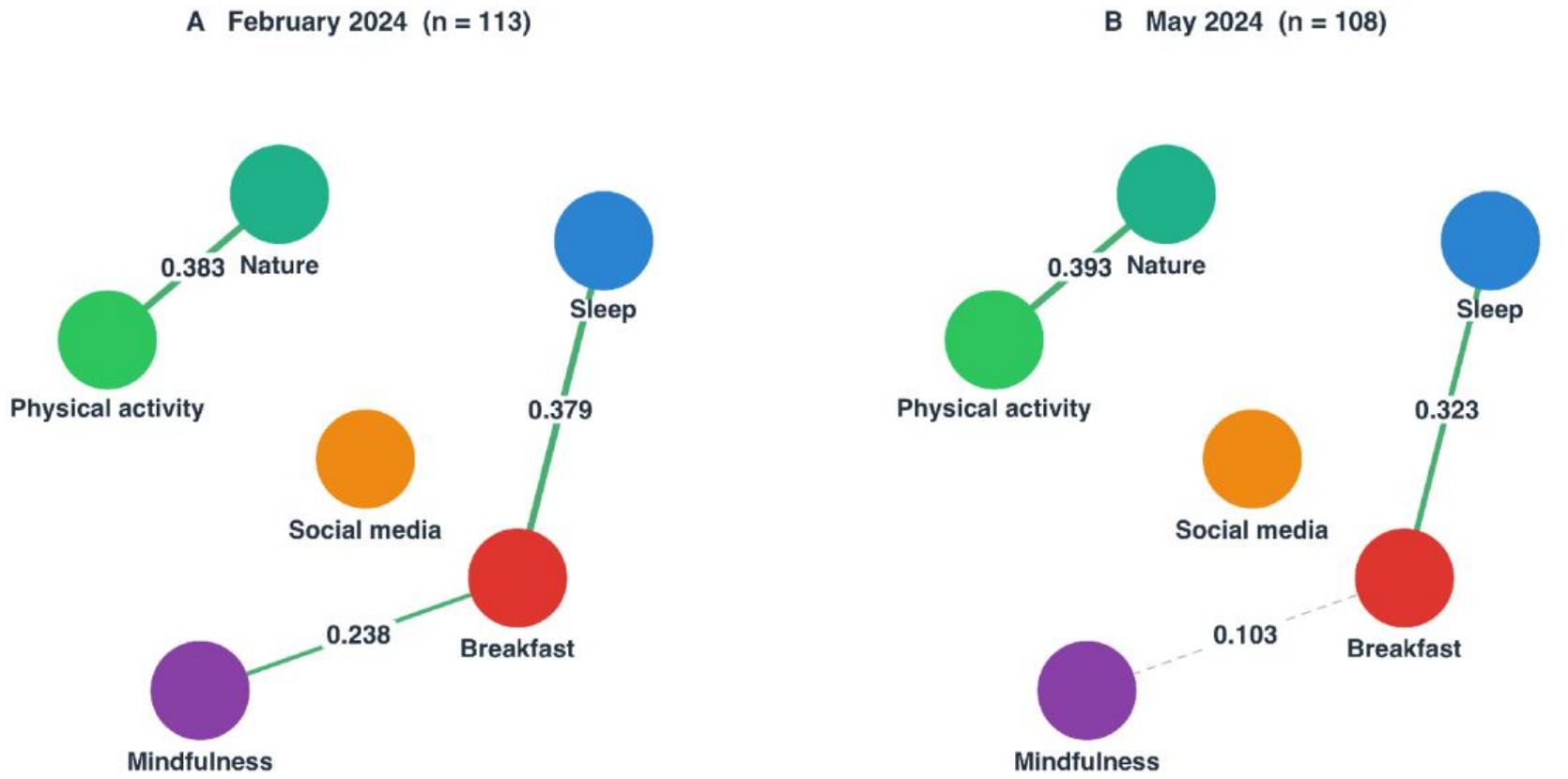
Contemporaneous health behavior correlation networks in Study 1. *Note*. Solid edges indicate |r| > .20. Positive edges are green; negative edges are muted red. Width is proportional to |r|; labels show Pearson correlations.

## 4 Study 2

### 4.1 Participants and procedure

For Study 2, we recruited 177 students at the same school using the same ethics and consent procedures as in Study 1. Data collection was conducted during September 23–26 2024 and again using the same procedures on May 9–30 2025. Of the 114 participants in Study 1, 74 also participated in Study 2, and 37 of those provided health behavior data at the Study 2 follow-up. The studies were not treated as independent samples or pooled for inference. At least one response on health behaviors was available for 176 students at Time 1 and 82 at Time 2. Complete data on all six behaviors were available for 170 students at Time 1 and 77 at Time 2, with 76 complete at both waves. Primary paired samples ranged from 77 to 82. Thus, 95 of the 177 records lacked all six Time 2 responses, and only 46.3% contributed any follow up information on health behaviors.

### 4.2 Results

None of the primary paired tests detected statistically significant change (Table 5). Unadjusted *p*-values were .635 for sleep, .469 for breakfast, .202 for physical activity, .812 for social media, .893 for mindfulness practices, and .365 for nature exposure. All *p*-values adjusted using the Holm method were 1.000. Physical activity showed the largest positive signed-rank effect (.201), while nature exposure showed a negative effect (-.157) (Table 2). Among the 76 complete participants, all tests remained nonsignificant (*p* = .108-.997).

**Table 5.**
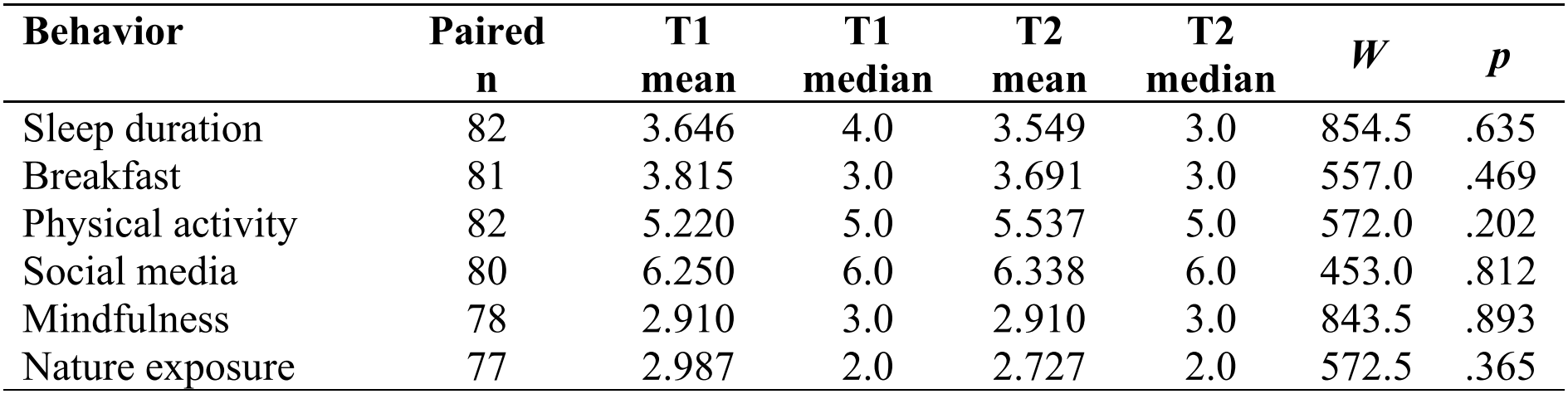
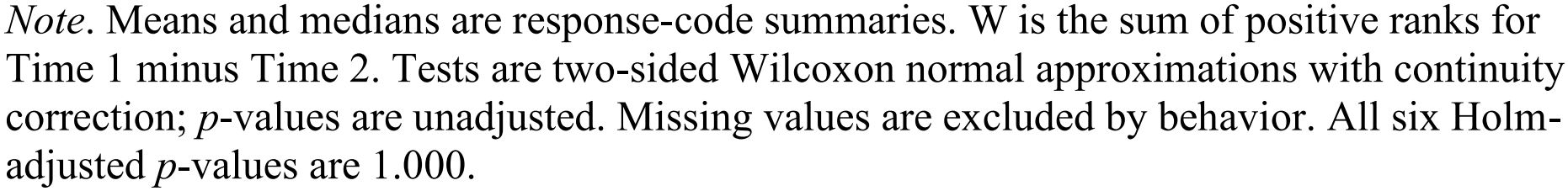
Paired health behavior comparisons in Study 2.

| **Behavior** | **Paired n** | **T1 mean** | **T1 median** | **T2 mean** | **T2 median** | ***W*** | ***p*** |
|---|---|---|---|---|---|---|---|
| Sleep duration | 82 | 3.646 | 4.0 | 3.549 | 3.0 | 854.5 | .635 |
| Breakfast | 81 | 3.815 | 3.0 | 3.691 | 3.0 | 557.0 | .469 |
| Physical activity | 82 | 5.220 | 5.0 | 5.537 | 5.0 | 572.0 | .202 |
| Social media | 80 | 6.250 | 6.0 | 6.338 | 6.0 | 453.0 | .812 |
| Mindfulness | 78 | 2.910 | 3.0 | 2.910 | 3.0 | 843.5 | .893 |
| Nature exposure | 77 | 2.987 | 2.0 | 2.727 | 2.0 | 572.5 | .365 |

*Note*. Means and medians are response-code summaries. W is the sum of positive ranks for Time 1 minus Time 2. Tests are two-sided Wilcoxon normal approximations with continuity correction; *p*-values are unadjusted. Missing values are excluded by behavior. All six Holm-adjusted *p*-values are 1.000.

The correlational pattern differed from Study 1. At Time 1, no Pearson correlation exceeded |r| = .20. Sleep-breakfast was close to the threshold (*r* = .193) (Table 3). At Time 2, five connections exceeded the display threshold: sleep-breakfast (.324), physical activity-nature (.268), sleep-social media (-.209), physical activity-social media (-.238), and mindfulness-nature (-.276; Figure 2; Table 3). Restricting the networks to the same 76 students preserved the broad pattern. No Time 1 Pearson edge exceeded .20, whereas the same five edges did at Time 2.

**Figure 2.**
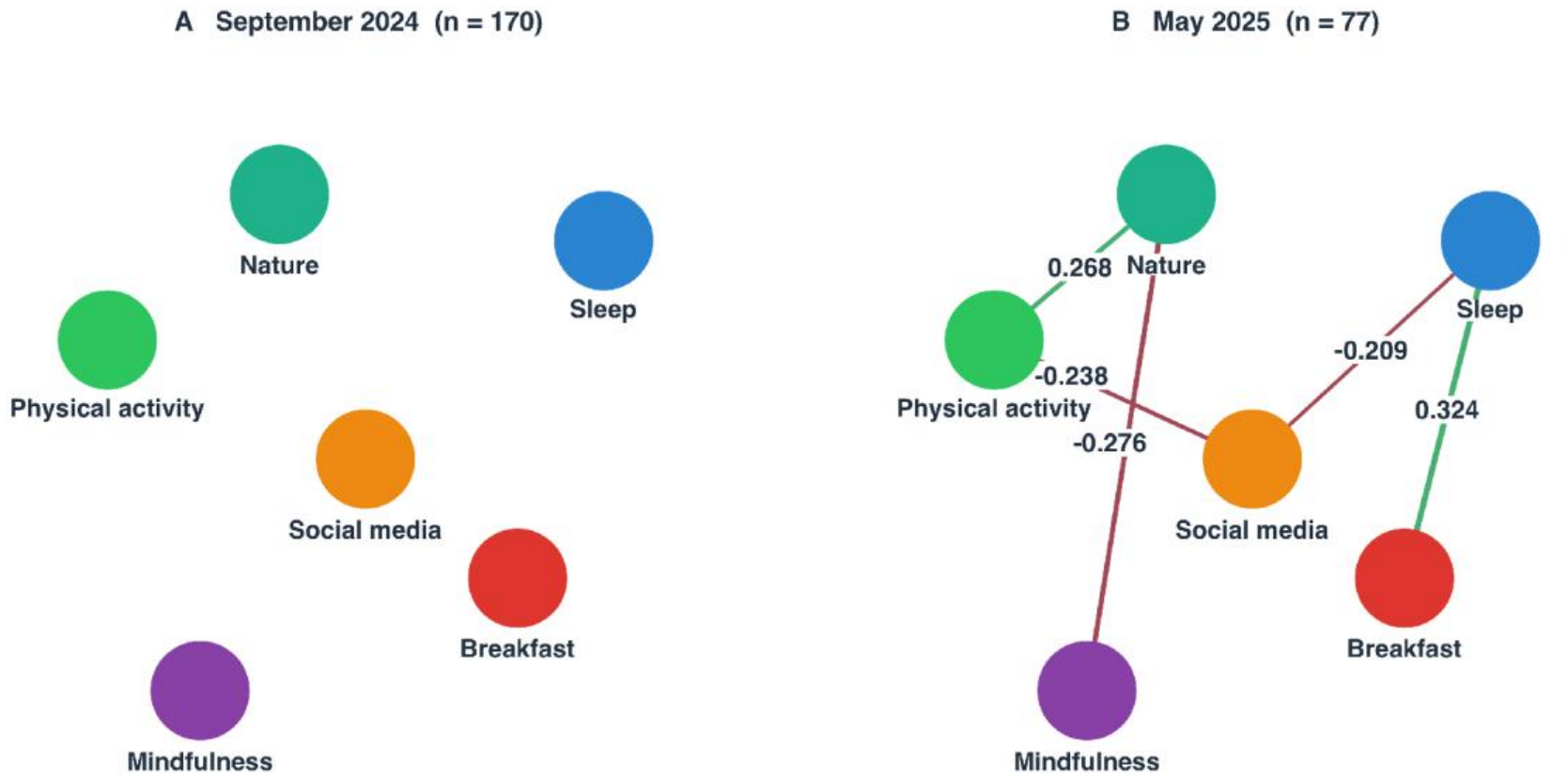
Contemporaneous health behavior correlation networks in Study 2. *Note*. Solid edges indicate |r| > .20. Positive edges are green; negative edges are muted red. Width is proportional to |r|; labels show Pearson correlations. No Time 1 correlation exceeded the cutoff. Networks use complete cases separately at each wave.

## 5 Discussion

### 5.1 Convergence in health behaviors

Across both studies, high school students showed no statistically significant group-level changes in physical activity or the other five assessed health behaviors. Study 1 covered the transition from late winter to late spring, whereas Study 2 extended from autumn to spring; thus, the convergence of the primary analyses across different intervals suggests limited detectable change in these samples. The physical activity findings are consistent with Rich et al.’s (2012) conclusion that seasonal effects on youth physical activity are not uniform across contexts. Their review showed that while UK-based studies commonly report higher activity in summer and lower in winter, several non-UK studies detect no seasonal differences. In addition to physical activity, all other health behaviors, including sleep duration, breakfast consumption, social media use, mindfulness practices, and nature exposure, also remained stable across the seasonal transition. This pattern of behavioral stability suggests that these habits function as enduring routines within this adolescent population. Structured school schedules and other contextual factors may have contributed to the observed patterns.

### 5.2 Recurring behavioral associations

Sleep-breakfast was the most consistently positive of the focal associations across the four assessments, although the Study 2 baseline coefficient fell just below .20. This pattern is compatible with research linking sleep health and breakfast consumption in adolescents (Lebacq *et al*., 2022; Mathew *et al*., 2022). Lebacq et al. (2022) reported that weekday sleep duration positively predicted daily breakfast consumption among over 8,000 adolescents, even after adjusting for morning tiredness. As pointed out by Mathew et al. (2022), poor sleep quality leads to rushed routines, increasing the likelihood of skipping breakfast. Accordingly, better sleep quality supports more consistent morning eating. This finding suggests that interventions targeting breakfast consumption might benefit from addressing sleep hygiene concurrently.

Physical activity and nature exposure were positively and prominently associated at three of the four assessments: both Study 1 waves and the Study 2 follow-up. This pattern is consistent with Bélanger et al. (2019), who found that outdoor time was associated with greater physical activity among youth. In contrast, the association was weak at the Study 2 baseline in September. One possible explanation is that September represents the beginning of the school year, when students may still be transitioning from less structured summer routines to more regular school schedules. Their physical activity and time in nature may therefore have occurred in varied and not necessarily overlapping contexts. For example, physical activity through organized sports or gym classes may have transitioned to indoor settings due to weather and school routine. This explanation remains tentative because the studies did not measure activity setting or adjustment to school routines.

Across the first three assessments, social media use showed no correlations above the network display threshold with the other health behaviors. At the May 2025 follow-up, however, social media use was negatively associated with sleep duration and physical activity, suggesting that its previously isolated position in the network did not persist. One possible explanation is that social media use became more frequent or more embedded in students’ daily routines by May 2025, increasing its potential to coincide with reduced sleep or physical activity. The present data cannot confirm this explanation, because the analyses could not establish that social media use caused changes in either behavior. Nevertheless, the emerging connections are relevant to Centola’s (2013) account of how social networks can shape health behaviors. Social media should be considered alongside other health behaviors in future assessments.

The breakfast-mindfulness connection observed in Study 1 did not recur above the display threshold in Study 2, suggesting that the original association may have been sample-specific. Measurement limitations may also have contributed. Both behaviors were assessed with single items, and the mindfulness practices measure captured only reported frequency of mindfulness practices. It did not distinguish the timing, type, duration, or quality of practices such as meditation, yoga, or breathing exercises. The Study 1 correlation may have reflected shared contextual factors rather than a direct relationship between breakfast and mindfulness practices. For example, students with more structured morning and evening routines, stronger family support, greater access to breakfast, or better overall self-regulation may have been more likely both to eat breakfast regularly and to practice mindfulness. Because these factors were not measured, the findings cannot determine whether either behavior influenced the other or whether both arose from the same routines and resources.

### 5.3 Implications for health education

For health education, the findings suggest that students’ reported behaviors were relatively stable at the group level across the observed intervals. For broad assessment of current group needs, a well-designed one-time survey may provide sufficient information for initial educational planning, particularly when repeated measurement is not feasible. The recurring sleep-breakfast association offers the clearest evidence for coordinated inquiry. The physical activity-nature association, which appeared at three assessments, similarly supports treating health behaviors as interconnected habits.

Social media use was isolated from the other health behaviors at the first three assessments, with no connections above the display threshold. At the Study 2 follow-up, however, it was negatively associated with sleep and physical activity, indicating that a concern that was not apparent at the earlier assessments had become more visible. These newly apparent connections have several practical implications for health education. First, digital-media education should be integrated with sleep education by addressing late-night use, notification management, and screen-free bedtime routines (Subramanian and Kumar, 2026; Williamson and Hale, 2024). Second, physical activity programs should address screen-based displacement by helping students identify periods of prolonged social media use and replace part of that time with accessible movement or outdoor activities (Melkevik *et al*., 2010). Third, educators should teach self-monitoring skills so students can examine how their own media-use patterns coincide with sleep duration, energy, and activity across the week. Fourth, school health curricula should present social media, sleep, and physical activity as interconnected components of daily routines rather than as separate topics, allowing students to develop realistic plans that balance digital participation with healthy behavior.

### 5.4 Priorities for future research

The present findings highlight several priorities for future research. First, future research should determine when a one-time assessment is sufficient for school health planning and when repeated assessment adds meaningful information. Studies could compare surveys administered at different points in the school year and test whether conclusions about group needs remain consistent across seasons and cohorts. Equivalence analyses using predefined thresholds for meaningful change would allow researchers to directly evaluate whether behaviors remain relatively stable.

Moreover, research should examine the mechanisms underlying the recurring and newly emerging behavioral connections. Intensive longitudinal designs, such as short daily diaries, could clarify the temporal relationships among sleep, breakfast, physical activity, nature exposure, and social media use. In particular, studies should investigate whether late-night social media use precedes shorter sleep, whether screen use displaces physical activity, and whether access to quality outdoor spaces such as parks and gardens helps explain the physical activity-nature association. Measuring timing, context, and barriers would move the evidence beyond cross-sectional correlations and identify specific processes that health promotion programming can address.

Lastly, future studies should test whether these patterns generalize across schools and can guide effective health education. Multi-school research should include diverse student populations, using consistent measures to distinguish broadly recurring associations from institution-specific patterns. Intervention studies could evaluate the effectiveness of integrated approaches, such as combining sleep and breakfast education or linking digital-media education with sleep and physical activity. Future studies should also capture various outcomes such as students’ health knowledge and well-being, school engagement, and academic performance. These areas of inquiry would show whether the observed associations can support practical educational decisions and measurable student benefits.

### 5.5 Strengths and limitations

This study has several strengths. It includes two longitudinal studies that used the same analytical approach, allowing the findings to be compared across study periods. Moreover, the assessments covered different parts of the school year. Study 1 followed students from late winter to late spring, while Study 2 followed students from autumn to spring. The analyses addressed two distinct yet related questions: changes in individual behaviors and associations among behaviors. They provided a broader view of behavioral stability and recurring patterns.

Several limitations should be considered. First, both studies were conducted at a single vocational-technical high school. The findings may reflect conditions specific to this school and may not generalize to other student populations or educational settings. Second, the number of paired observations was modest, especially in Study 2. Follow-up missingness was also substantial in Study 2, which reduced the amount of information available for evaluating change over time. Third, each health behavior was measured with a single ordinal item. The measures used different category widths and recall periods, and Pearson correlations treated the response codes as numerical values. Fourth, the analyses did not account for classroom clustering or participant characteristics. The inferential results assumed that students within each study were independent, yet there were some students who were surveyed in both studies. Fifth, the network graphs represented marginal correlations. They cannot identify causal pathways, changes within individuals, or intervention mechanisms. Finally, each study included only two assessment points. The data cannot describe complete seasonal trajectories, establish equivalence, or demonstrate statistically significant differences between the networks.

## 6. Conclusion

This study examined changes in six health behaviors and the recurrence of associations among those behaviors in vocational-technical high school students. It used two longitudinal studies conducted at the same school. Study 1 assessed students from late winter to late spring, while Study 2 assessed students from autumn to spring. Across these two studies, the six reported health behaviors showed relative stability at the group level. We did not find a significant shift in these behaviors across either assessment interval. For broad school health planning, a one-time survey may provide a useful picture of current group needs. Repeated assessment remains important when schools seek to evaluate longer-term behavioral patterns or the effects of a specific program. Sleep and breakfast showed the most consistent positive association, while physical activity and nature exposure were positively associated at three of the four assessments. These patterns support teaching related behaviors together within health education. Social media was isolated from the other behaviors at the first three assessments but was negatively associated with sleep and physical activity at the final assessment. This emerging pattern supports integrating digital-media education with sleep and physical activity education. Overall, the findings suggest that group-level behavior patterns may remain relatively stable even when relationships among behaviors change across assessments. Future research should test behavioral stability directly, clarify the mechanisms underlying recurring and emerging associations, and examine whether integrated health-education approaches produce measurable benefits across diverse school settings.

### Human Subjects Approval Statement

The study protocol was approved by the Lehigh University institutional IRB committee prior to data collection (2151549-5). Informed consent/assent was obtained for each participant, and parents were informed of the research and had an opportunity to opt their child out of participation.

## Data Availability

All data produced in the present study are available contingent upon IRB parameters and reasonable request to the authors

## Competing Interest Statement

All authors of this article declare they have no conflicts of interest. No authors or their institutions received any payments or services in the past 36 months from a third party that could be perceived to influence, or give the appearance of potentially influencing, the submitted work.

## Funding Statement

There were no sources of funding for the research.

## Notes

### Competing Interest Statement

The authors have declared no competing interest.

### Author Declarations

IRB of Lehigh University gave ethical approval for this work

## References

Bélanger, M., Gallant, F., Doré, I., O’Loughlin, J.L., Sylvestre, M.-P., Abi Nader, P., Larouche, R., et al. (2019), “Physical activity mediates the relationship between outdoor time and mental health”, Preventive Medicine Reports, Elsevier, Vol. 16, p. 101006.

Bucher KA, Moriarty CM, Lazarchak A, McIntire RK. Teachers’ Perspectives on the Impact of an Outdoor-Based Self-Care Training Program on Student Mental Health. Int J Environ Res Public Health. 2025;22(7):1135.

Centola, D. (2013), “Social media and the science of health behavior”, Circulation, Lippincott Williams & Wilkins Hagerstown, MD, Vol. 127 No. 21, pp. 2135–2144.

Gignac, G.E. and Szodorai, E.T. (2016), “Effect size guidelines for individual differences researchers”, Personality and Individual Differences, Elsevier, Vol. 102, pp. 74–78.

Harris, K.M. (2010), “An integrative approach to health”, Demography, Springer, Vol. 47 No. 1, pp. 1–22.

Hevey, D. (2018), “Network analysis: a brief overview and tutorial”, Health Psychology and Behavioral Medicine, Taylor & Francis, Vol. 6 No. 1, pp. 301–328.

Hjorth, M.F., Chaput, J.-P., Michaelsen, K., Astrup, A., Tetens, I. and Sjödin, A. (2013), “Seasonal variation in objectively measured physical activity, sedentary time, cardio-respiratory fitness and sleep duration among 8–11 year-old Danish children: a repeated-measures study”, BMC Public Health, Springer, Vol. 13 No. 1, p. 808.

Lebacq, T., Holmberg, E., Pedroni, C., Dujeu, M. and Castetbon, K. (2022), “Weekday sleep duration and morning tiredness are independent covariates of breakfast skipping in adolescents”, European Journal of Clinical Nutrition, Nature Publishing Group UK London, Vol. 76 No. 10, pp. 1403–1408.

Mathew, G.M., Reichenberger, D.A., Master, L., Buxton, O.M., Hale, L. and Chang, A.-M. (2022), “Worse sleep health predicts less frequent breakfast consumption among adolescents in a micro-longitudinal analysis”, International Journal of Behavioral Nutrition and Physical Activity, Springer, Vol. 19 No. 1, p. 70.

Melkevik, O., Torsheim, T., Iannotti, R.J. and Wold, B. (2010), “Is spending time in screen-based sedentary behaviors associated with less physical activity: a cross national investigation”, International Journal of Behavioral Nutrition and Physical Activity, Springer, Vol. 7 No. 1, p. 46.

Rich, C., Griffiths, L.J. and Dezateux, C. (2012), “Seasonal variation in accelerometer-determined sedentary behaviour and physical activity in children: a review”, International Journal of Behavioral Nutrition and Physical Activity, Springer, Vol. 9 No. 1, p. 49.

Subramanian, R.K. and Kumar, D. (2026), “Effect of a sleep hygiene-focused health education intervention on screen-time behaviours and Pittsburgh sleep quality index outcomes”, Journal of Chemical Health Risks, Vol. 16 No. 3, p. 1.

Tamanal, J.M. and Kim, C.H. (2020), “Promoting healthy lifestyle in high school students: Determination of the lifestyle status through the healthy lifestyle screen (HLS) assessment”, Journal of Lifestyle Medicine, Vol. 10 No. 1, p. 30.

Williamson, A.A. and Hale, L. (2024), “Future directions for screen time interventions for sleep”, JAMA Pediatrics, Vol. 178 No. 12, pp. 1245–1247.

